# SARS-CoV-2 seroprevalence in Delhi, India - September-October 2021 – a population based seroepidemiological study

**DOI:** 10.1101/2021.12.28.21268451

**Authors:** Pragya Sharma, Saurav Basu, Suruchi Mishra, Ekta Gupta, Reshu Aggarwal, Pratibha Kale, Nutan Mundeja, B S Charan, Gautam Kumar Singh, Mongjam Meghachandra Singh

**Affiliations:** Maulana Azad Medical College, New Delhi; Indian Institute of Public Health – Delhi; Institute of Liver and Biliary Sciences, New Delhi; Directorate General Health Services, Government of NCT, Delhi

**Keywords:** Serosurvey, Seroprevalence, Covid-19, Vaccination, India

## Abstract

**Background:** We conducted a repeat serosurvey in Delhi, India to estimate the seroprevalence of SARS-CoV-2 in the general population and compare the antibody prevalence in the vaccinated and non-vaccinated groups.

**Methods:** This cross-sectional study was conducted from September 24 to October 14 2021 in 280 wards of Delhi among 27811 participants selected through a multistage sampling technique with housing settlement based stratification. The SARS-CoV-2 immunoglobulin (IgG) antibodies were screened with the VITROS® (Ortho Clinical Diagnostics, Raritan, NJ, USA) assay (90% sensitivity, 100% specificity).

**Results:** A total of 24895 (89.5%) samples were seropositive. The crude seroprevalence was 87.99% (95% CI 89.1, 89.8), weighted for age and sex was 88% (95% CI 87.6, 88.4), and after adjustment of assay performance was estimated as 97.5% (95% CI 97.0, 98.0). The weighted seroprevalence in the 11 districts ranged from 84.9% (South-West district) to 90.8% (East district) Females in all the age-groups (<18, 18-49 and ≥50) had significantly higher odds of seropositivity (p<0.001). On adjusted analysis, the odds of seroconversion in the participants vaccinated with at-least one dose of either Covid-19 vaccine (Covishield/Covaxin) was more than four times compared to the unvaccinated (aRR 4.2 (3.8, 4.6)). The seroprevalence was also comparable among the complete and partially vaccinated subgroups for both vaccines (Table 4). Most (86.8%) seropositive individuals had a SARS-CoV-2 signal/cut-off ≥4.0 except in children

**Conclusions:** We observed IgG antibodies against SARS-CoV-2 in most of the general population of Delhi with likely higher antibody titres in the vaccinated compared to the unvaccinated groups.

## INTRODUCTION

Delhi, the capital city-state of India, experienced a severe second wave of Covid-19, predominantly driven by the Delta variant during April-June 2021 [1, 2]. The city having a population of ∼19 million, recorded ∼1.43 million cases and 25090 deaths until October 21 2021 [3].

Covid-19 seroprevalence studies estimate the population level humoral immunity profile by direct measurement of SARS-CoV-2 antibodies induced through either natural infection or vaccination [4]. Repeated cross-sectional seroepidemiological studies when regionally localized enable seroprevalence monitoring for guiding infection prevention and control strategies by mounting effective public health interventions [5].

Serial serosurveys previously indicated that the age and sex weighted seroprevalence in Delhi in the ≥5 population increased from 24.71% (95% CI 24.01 to 25.42) in August-October 2020 to 50.52% (95% CI 49.94 to 51.10 in January 2021 [6, 7].

India’s Covid-19 vaccination campaign was formally initiated from January 16 2021 and predominantly driven with ChAdOx1 nCoV-19 (Covishield, Serum Institute of India, Pune) and BBV152 (Covaxin; Bharat Biotech International, Hyderabad). The vaccination strategy initially restricted to healthcare and frontline workers (January 16 2021 onwards) was expanded initially for elderly (>60 years old) and select comorbid (>45 years old) individuals (March 1 2021 onwards), all >45 years (April 1 2021 onwards), and later all individuals >18 years old (May 1 2021 onwards). Vaccination has not yet been initiated in children (<18 years) till date [8]. As of 21st October 2021, 19.8 million cumulative vaccine doses were administered to the eligible beneficiaries [8].

We conducted this sixth round of Delhi state serosurvey to estimate the seroprevalence of SARS-CoV-2 in the general population and compare the antibody prevalence in the vaccinated and non-vaccinated groups.

## METHODS

### Study design, participants, and settings

This was a cross-sectional, seroepidemiological study among individuals aged five and above who were recruited from 274 wards in the state of Delhi from September 24 to October 14 2021.

A total of 100 participants each were enrolled from all wards except the Delhi Cantonment and the New Delhi wards. The sample size of approximately 28,000 was estimated at 99% confidence level, 1% absolute precision, 56% expected prevalence from the previous serosurvey [7], design effect of 1.5, and considering a non-response rate of 10%.

The sampling was conducted within each ward based on the residential settlement types which included planned colonies, urban slums, resettlement colonies, unauthorized colonies, and rural areas [9]. Within each ward, the proportion of participants selected from each settlement type was stratified according to their tentatively estimated population size. The participants were selected for this household serosurvey through a multistage sampling technique having the following steps: (i). Sampling areas within each settlement type by simple random sampling (ii) Household selection by systematic random sampling, and (iii). Age-order procedure for selection of a single individual within each selected household.

### Laboratory procedure

A trained phlebotomist or lab technician under aseptic precautions collected 3-4 mL of venous blood which were transported and processed at the designated laboratory. Anti-SARS-CoV-2 IgG antibodies were detected using chemiluminescent technology based VITROS® assay on VITROS® 3600 (Ortho Clinical Diagnostics, Raritan, NJ, USA) [10]. The assay has a documented sensitivity 90% and specificity of 100% which is deemed acceptable for SARS-CoV-2 seroprevalence surveys [11].

### Statistical analysis

A customized android tablet application was used for electronic data collection by field volunteers recruited for this survey. The variables included sociodemographic data, past history of Covid-19 disease, and Covid-19 vaccination status. This questionnaire data was subsequently merged with the laboratory antibody test result data in Microsoft Excel 2013. The data were analysed using SPSS Version 25 (IBM Corp., Armonk, NY, USA). The seroprevalence estimates were weighted to match the state demographics by age and sex and reported as proportions with 95% confidence intervals (CIs). The adjusted seroprevalence was estimated through statistical correction of the weighted seroprevalence by incorporating the assay characteristics in the Rogan-Gladen estimator, where true (adjusted) prevalence = weighted prevalence + (specificity - 1)/(specificity + sensitivity - 1) [12]. Results were expressed as frequency and proportions for categorical variables and mean and standard deviation for continuous variables. The chi-square test was used to assess association between categorical variables. A binary logistic regression analysis was conducted by considering IgG antibody as the outcome and the following independent variables; sex: male/female, age: <18/≥18, settlement: planned/other, past-history of Covid-19: present/absent, and vaccination status: no dose/at-least one dose. A p-value of <0.05 was considered statistically significant.

### Ethics

The study was approved by the Institutional Ethics Committee, Maulana Azad Medical College & Associated Hospitals, New Delhi vide F.1/IEC/MAMC/85/03/2021/No428 dated 21.08.2021. Electronic and informed consent was obtained from all the study participants.

## RESULTS

A total of 28491 laboratory samples were collected of which 27811 were successfully processed in the laboratories (Table 1).

**Table 1.** Seroprevalence of antibodies to SARS-CoV-2 across districts of Delhi, September-October 2021

| District | Seroprevalence (Crude) | Seroprevalence (Weighted) |
| --- | --- | --- |
| Central | 91.2 | 90.5 |
| East | 91.7 | 90.8 |
| New Delhi | 88.4 | 87.0 |
| Shahdara | 88.3 | 85.8 |
| North | 87.9 | 86.5 |
| NED | 92.0 | 90.7 |
| NWD | 90.7 | 89.6 |
| South | 90.6 | 90.0 |
| SED | 89.1 | 87.2 |
| SWD | 86.7 | 84.9 |
| West | 89.1 | 87.5 |

A total of 24895 (89.5%) samples were seropositive. The crude seroprevalence was 87.99% (95% CI 89.1, 89.8). The seroprevalence weighted for age and sex was 88% (95% CI 87.6, 88.4). The weighted seroprevalence in the districts ranged from 84.9% (South-West district) to 90.8% (East district) (Table 1). The adjusted seroprevalence, calculated after statistical correction the weighted seroprevalence for assay characteristics was 97.5% (95% CI 97.0, 98.0). On bivariate analysis, females in all the age-groups (<18, 18-49 and ≥50) had significantly higher odds of seropositivity (p<0.001) (Table 2).

**Table 2.** Age and sex stratified seroprevalence of antibodies to SARS-CoV-2, Delhi, September-October 2021

| Sex | Total (n=27811) | IgG Seropositive | IgG Seronegative | Adjusted odds (95% C.I) |
| --- | --- | --- | --- | --- |
| Male |  |  |  |  |
| <18 | 2165 (18.4) | 1765 (81.5) | 400 (18.5) | 1 (p<0.001) |
| 18-49 | 6838 (58.0) | 6147 (89.9) | 691 (10.1) | 2.0 (1.76, 2.3) |
| ≥50 | 2788 (23.6) | 2489 (89.3) | 299 (10.7) | 1.88 (1.6, 2.21) |
| Female |  |  |  |  |
| <18 | 2046 (12.8) | 1680 (82.1) | 366 (17.9) | 1 (p<0.001) |
| 18-49 | 10357 (64.7) | 9388 (90.6) | 969 (9.4) | 2.11 (1.85, 2.40) |
| ≥50 | 3617 (22.5) | 3426 (94.7) | 191 (5.3) | 3.9 (3.25, 4.7) |

Our logistic regression model was statistically significant with Chi2(5) = 1105.5 (p<0.001) and correctly classified 89.3 % of the cases. The Hosmer Lemeshow goodness of fit test-statistic had p value of 0.124 from which we concluded that the model estimates the data acceptably. Based on this adjusted analysis, we found the participants of female gender (aRR 1.3 [1.2, 1.4]), and those having received at-least one dose of either Covid-19 vaccine (aRR 4.2 (3.8, 4.6) as statistically significant predictors of SARS-CoV-2 seropositivity (p<0.001) (Table 3). The seroprevalence was also comparable among the complete and partially vaccinated subgroups for both vaccines (Table 4).

**Table 3.**
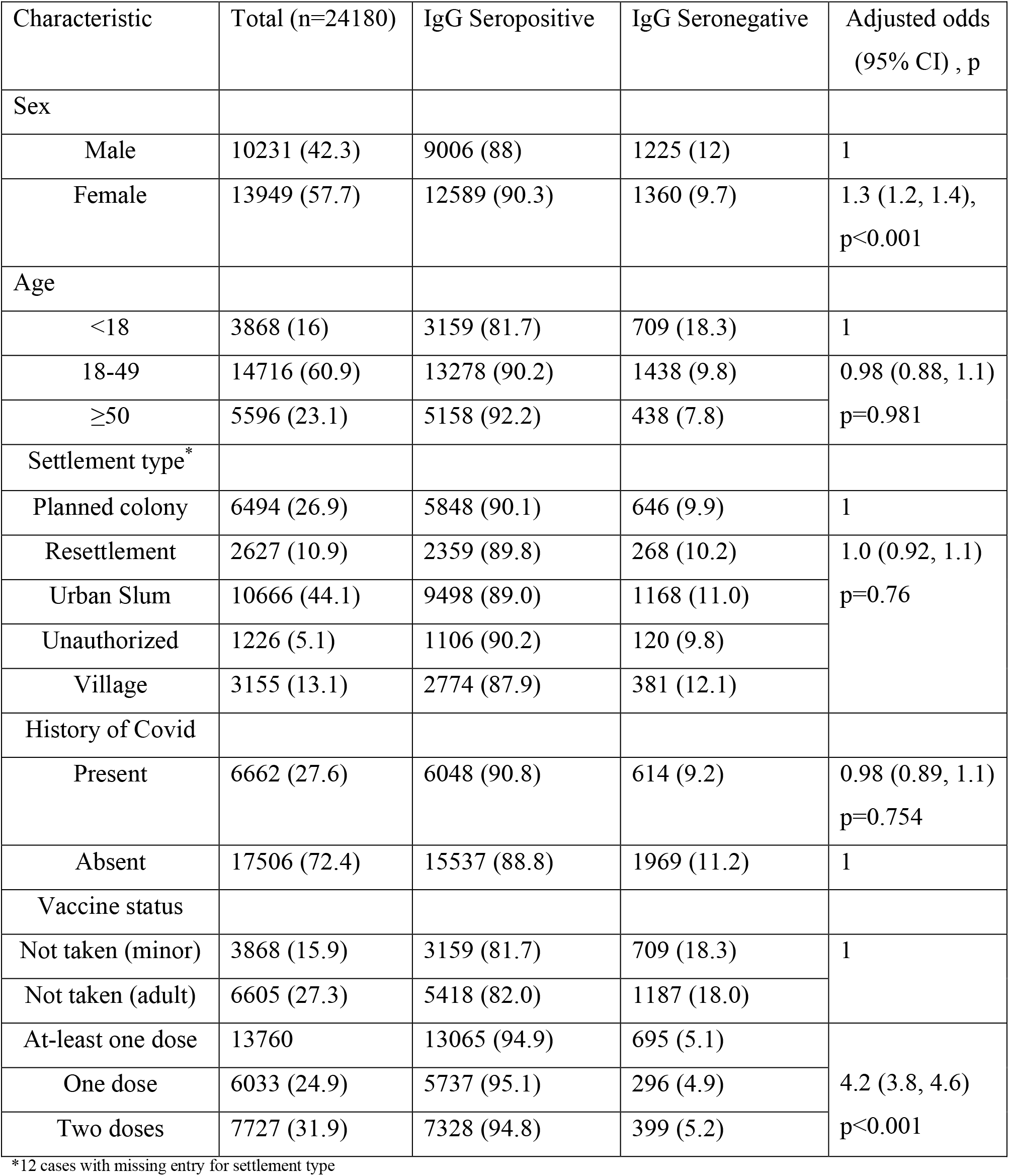
Seroprevalence of antibodies to SARS-CoV-2, Delhi, September-October 2021

**Table 4.** Vaccination status and seroprevalence of antibodies to SARS-CoV-2, Delhi, September-October 2021*

| Vaccine type* | Total (n=13760) | IgG Seropositive (95% C.I.) |
| --- | --- | --- |
| Covishield 1 dose | 5140 (37.3) | 4914 (95.6) (95.0, 96.1) |
| Covishield 2 doses | 6086 (44.2) | 5802 (95.3) (94.8, 95.8) |
| Covaxin 1 dose | 889 (6.5) | 819 (92.1) (90.2, 93.7) |
| Covaxin 2 doses | 1628 (11.8) | 1514 (93.0) (91.6, 94.1) |
| History of Covid-19 present (n=6662) |  |  |
| Covishield 1 dose | 1603 (24.1) | 1541 (96.1) (95.1, 97.0) |
| Covishield 2 doses | 2095 (31.4) | 2006 (95.8) (94.8, 96.5) |
| Covaxin 1 dose | 259 (3.9) | 238 (91.9) (87.9, 94.6) |
| Covaxin 2 doses | 554 (8.3) | 516 (93.1) (90.7, 95.0) |
| No vaccination | 2143 (32.2) | 1739 (81.1) (79.4, 82.7) |
| No history of Covid-19 (n=17506) |  |  |
| Covishield 1 dose | 3537 (20.2) | 3373 (95.4) (94.6, 96.0) |
| Covishield 2 doses | 3991 (22.8) | 3796 (95.1) (94.4, 95.7) |
| Covaxin 1 dose | 630 (3.6) | 581 (92.2) (89.9, 94.1) |
| Covaxin 2 doses | 1074 (6.1) | 998 (92.9) (91.2, 94.3) |
| No vaccination | 8265 (47.2) | 6781 (82.0) (81.2, 82.9) |
| Diabetes Mellitus (n=1273) |  |  |
| Covishield 1 dose | 219 (17.2) | 215 (98.2) (95.4, 99.3) |
| Covishield 2 doses | 476 (37.4) | 450 (94.5) (92.1, 96.2) |
| Covaxin 1 dose | 70 (5.5) | 68 (97.1) (90.2, 99.2) |
| Covishield 2 doses | 174 (13.7) | 158 (90.8) (85.6, 94.3) |
| No vaccination | 313 (24.6) | 262 (83.7) (79.2, 87.4) |

\*A total of 13 participants had taken the Sputnik vaccine

The signal/ cut off (S/CO) of SARS-CoV-2 IgG in the seropositive samples (n=24,895) ranged from 1.00 to 22.8 (median 11.40, IQR 6.97, 14.5). A total of 21620 (86.8%) seropositive individuals had a S/CO ≥4.0, signifying the presence of high antibody titres. The S/CO <4 was observed in 14.2% female seropositive participants compared to 12.4% male seropositive participants, and this difference was statistically significant (p<0.001). Furthermore, S/CO <4 was observed in 29.1% participants aged below 18 compared to 11.1% and 9.2% in the 18-49 and ≥50 age-group seropositive participants, respectively (p<0.001). The robust antibody response translated into effective population protection. From August 1 2021 to October 26 2021, the state on average daily recorded 39 new cases (range 0 to 151), and 0.44 deaths (range 0 to 5).

## DISCUSSION

The findings of the sixth round of the serosurvey in Delhi indicate that nearly nine in ten individuals aged 5 years and above in Delhi had detectable IgG SARS-CoV-2 antibodies which on assay adjustment reflect near universal seropositivity. The seroprevalence in Delhi was higher compared to other Indian cities and states but comparable with Mumbai city (86.64%) probably because of the high severity of the second wave of the pandemic in these metropolitan cities (Table 5).

**Table 5.** SARS-CoV-2 Seroprevalence in Indian states and cities after second wave of Covid-19 pandemic

| SN | Site | Area | Age group | Sample Size | Study Duration | (Seropositivity) (%) |
| --- | --- | --- | --- | --- | --- | --- |
| 1. | Mumbai ( 5<sup>th</sup> Serosurvey) | 24 wards | >18 years | 8,674 | 8<sup>th</sup> August – 9<sup>th</sup> September | 86.64% |
| 2. | Tamil Nadu ( 3<sup>rd</sup> Serosurvey) | 37 districts (827 Clusters) |  | 24,586 | July and August | 70% |
| 3. | Punjab Health Department |  | 6-17 years | 1,577 |  | 58% |
| 4. | Kerala Health Department |  | >18 years | 13,000 | September | >18 Years: 82.6%<br>5-17 years: 40% |
| 5. | Himachal Pradesh | 12 districts | >6 years | 4,822 |  | 87.5% |
| 6. | Kashmir | 10 districts | Police personnel, health care workers, pregnant women | 5,663 | July | 84% |
| 7. | Haryana (3<sup>rd</sup> Serosurvey) | 22 districts | >6 years | 36,520 | September | 76.3% |
| 8. | Gujarat |  | - | 5,001 | 28 May- June 3rd | 81.63% |

1. https://timesofindia.indiatimes.com/city/mumbai/covid-19-mumbais-fifth-sero-survey-shows-prevalence-of-antibodies-at-86-64-per-cent/articleshow/86293984.cms
2. https://www.hindustantimes.com/india-news/tn-sero-survey-shows-70-have-antibodies-against-covid19-101633719330153.html
3. https://www.tribuneindia.com/news/patiala/58-children-in-punjab-have-antibodies-finds-sero-survey-296362
4. https://indianexpress.com/article/cities/thiruvananthapuram/coronavirus-82-per-cent-in-18-age-group-have-antibodies-finds-kerala-sero-survey-7566955/
5. https://www.hindustantimes.com/cities/chandigarh-news/87-himachal-population-has-covid-antibodies-sero-survey-101632509353759.html
6. https://www.hindustantimes.com/cities/others/sero-survey-covid-antibodies-found-in-84-of-kashmir-s-population-101631218975811.html
7. http://timesofindia.indiatimes.com/articleshow/87121689.cms?utm_source=contentofinterest&utm_medium=text&utm_campaign=cppst
8. https://www.hindustantimes.com/india-news/over-81-people-in-ahmedabad-have-covid-19-antibodies-serological-survey-finds-101626707621460.html

The weighted and adjusted seroprevalence of IgG SARS-CoV-2 antibodies in Delhi increased from 50.1% and 56.1% in January 2021 to 88.1% and 97%, respectively during September-October 2021 [7]. During the second wave of the countrywide Covid-19 pandemic, Delhi recorded nearly 0.737 million cases including 11075 deaths [2]. Consequently, like the national pattern, the increase in seroprevalence was due to both natural infection and vaccination [13]. However, during this nine-month period, the seroprevalence increased in unvaccinated adults from 50.3% to 82%, and in children aged five years and above from 52.4% to 81.7%, which is indicative of natural infection being the primary driver of immune response in a large proportion of the population.

Similar to the trends in previous serosurvey rounds, the seroprevalence was higher in adults compared to children and adolescents, in those having past-history of Covid-19, and in residents of planned colonies in comparison to other settlements. The seropositivity was also significantly higher in females compared to males despite lower rates of vaccination in the former suggesting a biological rather than behavioural phenomenon [6, 7].

Previous studies suggest antibodies to SARS-CoV-2 persist at-least one year after natural infection conferring durable protection against reinfection or symptomatic disease [14]. Unlike a previous study, we found only a small difference in the rates of seroconversion between the Covishield and Covaxin vaccines [15]. Furthermore, the proportion of seronegative vaccinated individuals in our study was very low even in those who had received only one dose of either vaccine. High seroprevalence was observed even in DM patients despite the possible association with an immunocompromised condition. However, in comparison, the nationwide ICMR study reported 10-14% of the vaccinated individuals remained seronegative even after receiving two vaccine doses [13]. This is possible because of the comparatively higher burden of natural infection in Delhi throughout the pandemic and later during second wave, and the fact that even a single dose of Covid-19 vaccination post-infection is known to induce robust antibody response [16, 17].

High S/CO suggestive of high antibody titres were observed in more than five of every six seropositive participants, while comparatively lower titres in the under-18 age group were likely because of their absence of vaccination. These findings indicate that a majority of the adults in Delhi have developed hybrid immunity due to Covid-19 vaccination after natural infection which is known to produce robust immune response [18]. The small number of average daily Covid-19 cases and very low-test positivity rate after the subsiding of the second wave in Delhi is suggestive of the attainment of the herd immunity threshold in the population with the caveat for the expected waning of antibody titres over time [19, 20].

The study has certain limitations. First, due to duplicate unique id generated by field data enumerators and failure of app validated check, questionnaire data of ∼10% participants could not be matched with the unique identification labels of their laboratory samples resulting in a loss of their sociodemographic (except age and sex recovered from laboratory data) and vaccination attributes. During analysis, this missing data was assumed to be missing at random type. For a similar reason, the exact response rate of the survey could not be established although based on consultation with field nodal officers, it was not dissimilar from the previous round (<20%). Second, in this investigation, we could not differentiate between vaccination or natural infection induced antibody response since both anti-S and anti-N antibodies are generated by inactivated vaccines such as Covaxin [11]. Third, the S/CO only provided an indirect marker of IgG SARS-CoV-2 antibody titres and was not an absolute correlate of immunological protection. Moreover, immune protection against SARS-CoV-2 can be both triggered through both humoral and cell media immunity, and consequently, seropositivity may not necessarily be indicative of protection against Covid-19 disease or vice versa [11, 21]. Finally, the participant’s vaccination status and Covid-19 disease history were often ascertained from individual recall in absence of validation with the vaccination certificate and laboratory record, respectively.

In conclusion, the findings of this serosurvey indicate the presence of IgG antibodies against SARS-CoV-2 in most of the general population of Delhi, although there exists a statistically significant gap in seroprevalence between the vaccinated and the unvaccinated groups. Consequently, vaccination coverage needs acceleration in the unvaccinated and the partially vaccinated adults especially for protection against severe disease and death. The emergence of the Omicron variant of concern which has been reported to have the ability to bypass both natural and vaccine induced immunity has major ramifications for the public health surveillance and response systems [22]. Nevertheless, the presence of hybrid immunity in a majority of the adult population in Delhi and India may confer enhanced protection from symptomatic or severe disease in comparison to populations in whom vaccine induced immunity is predominant. Continuous genomic surveillance, pandemic preparedness, and adherence to non-pharmaceutical interventions without any laxity or complacence requires high and sustained prioritization to diminish morbidity and mortality from future waves of the pandemic.

## Data Availability

All data produced in the present study are available upon reasonable request to the authors

## Conflicts of interest

None

## Sources of funding

This research received no specific funding from any agency in the public, commercial or not-for-profit sectors. The logistics and human resources were deputed by the Directorate General of Health Services, government of the National Capital Territory, Delhi and supported by the ATE Chandra Foundation and ACT grants.

## Acknowledgments

We thank all the district Nodal officers of Delhi for facilitating the data and sample collection. We express thanks to the ATE Chandra Foundation and ACT grants, and IDFC Foundation for technical support. We also thank Ms. Arti Kakkar for her assistance with data management for this investigation.

## Notes

### Competing Interest Statement

The authors have declared no competing interest.

### Author Declarations

The study was approved by the Institutional Ethics Committee, Maulana Azad Medical College & Associated Hospitals, New Delhi vide F.1/IEC/MAMC/85/03/2021/No428 dated 21.08.2021

## REFERENCES

1. Government of India. #IndiaFightsCorona COVID-19. New Delhi: Government of India; 2021. Available from: https://www.mygov.in/covid-19. (Accessed 15th December 2021)

2. Delhi reported more COVID cases, deaths in April-May than since beginning of pandemic. (2021). Available from: https://www.news18.com/news/india/delhi-reports-more-covid-cases-deaths-in-april-may-than-since-the-beginning-of-pandemic-3751346.html (Accessed 15th December 2021)

3. Government of India Ministry of Health and Family Welfare. Covid-19 dashboard. Available from: mohfw.gov.in (Accessed 22nd October 2021)

4. Alter G, Seder R. The power of antibody-based surveillance. N Engl J Med. 2020;383(18):1782–4

5. WHO. Interpreting SARS-CoV-2 seroprevalence studies for public health decision-making: Operational Brief. 7th December 2021

6. Sharma N, Sharma P, Basu S, Saxena S, Chawla R, Dushyant K, et al. The seroprevalence of severe acute respiratory syndrome coronavirus 2 in Delhi, India: a repeated population-based seroepidemiological study. Trans R Soc Trop Med Hyg. 2021 Aug 2:trab109. doi: 10.1093/trstmh/trab109.

7. Sharma N, Sharma P, Basu S, Bakshi R, Gupta E, Agarwal R, et al. Second Wave of the COVID-19 Pandemic in Delhi, India: High Seroprevalence Not a Deterrent? Cureus. 2021 Oct 24;13(10):e19000. doi: 10.7759/cureus.19000.

8. Ministry of Health and Family Welfare. Frequently asked questions. New Delhi: Ministry of Health and Family Welfare; 2021. Available from: https://www.mohfw.gov.in/covid_vaccination/vaccination/faqs.htm (Accessed 15th December 2021)

9. Centre for Policy Research. Categorisation of settlement in Delhi. 2015. Available from: https://www.cprindia.org/sites/default/files/policy-briefs/Categorisation-of-Settlement-in-Delhi.pdf (Accessed 15th December 2021)

10. Instructions for use - CoV2G. VITROS. 2021 Available from: https://www.fda.gov/media/137363/download 2021 (Accessed 15th December 2021)

11. World Health Organization. COVID-19 Target product profiles for priority diagnostics to support response to the COVID-19 pandemic v.1.0. 2020 Available from: https://www.who.int/publications/m/item/covid-19-target-product-profiles-for-priority-diagnostics-to-support-response-to-the-covid-19-pandemic-v.0.1 (Accessed 15th December 2021)

12. Estimating prevalence from the results of a screening test. Rogan WJ, Gladen B. Am J Epidemiol. 1978;107:71–76

13. Murhekar MV, Bhatnagar T, Thangaraj JWV, Saravanakumar V, Santhosh Kumar M, Selvaraju S, et al. Seroprevalence of IgG antibodies against SARS-CoV-2 among the general population and healthcare workers in India, June-July 2021: A population-based cross-sectional study. PLoS Med. 2021;18(12):e1003877.

14. Hall VJ, Foulkes S, Charlett A, Atti A, Monk EJM, Simmons R, et al. SARS-CoV-2 infection rates of antibody-positive compared with antibody-negative health-care workers in England: a large, multicentre,prospective cohort study (SIREN). Lancet. 2021; 397:1459–69

15. Singh AK, Phatak SR, Singh R, Bhattacharjee K, Singh NK, Gupta A, et al. Antibody response after first and second-dose of ChAdOx1-nCOV (CovishieldTM®) and BBV-152 (CovaxinTM®) among health care workers in India: The final results of cross-sectional coronavirus vaccine-induced antibody titre (COVAT) study. Vaccine. 2021;39(44):6492–6509.

16. Jamiruddin R, Haq A, Khondoker MU, Ali T, Md. Fa, Khandker SS, et al. Antibody response to the first dose of AZD1222 vaccine in COVID-19 convalescent and uninfected individuals in Bangladesh. Expert Rev Vaccines. 2021;20(12):1651–1660.

17. Eyre DW, Lumley SF, O’Donnell D, Stoesser NE, Matthews PC, Howarth A, et al. Stringent thresholds in SARS-CoV-2 IgG assays lead to under-detection of mild infections. BMC Infect Dis. 2021;21(1):187.

18. Parai D, Choudhary HR, Dash GC, Sahoo SK, Pattnaik M, Rout UK, et al. Single-dose of BBV-152 and CHADOX1 NCOV-19 increases antibodies against spike glycoprotein among healthcare workers recovered from SARS-CoV-2 infection. Travel Med Infect Dis. 2021;44:102170.

19. Kojima N, Klausner JD. Protective immunity after recovery from SARS-CoV-2 infection. Lancet Infect Dis. 2021;S1473-3099(21)00676-9

20. Lumley SF, Wei J, O’Donnell D, Stoesser NE, Matthews PC, Howarth A, et al. The Duration, Dynamics, and Determinants of Severe Acute Respiratory Syndrome Coronavirus 2 (SARS-CoV-2) Antibody Responses in Individual Healthcare Workers. Clin Infect Dis. 2021;73(3):e699–e709.

21. Cox RJ, Brokstad KA. Not just antibodies: B cells and T cells mediate immunity to COVID-19. Nat Rev Immunol. 2020 Oct;20(10):581–582.

22. Ferré VM, Peiffer-Smadja N, Visseaux B, Descamps D, Ghosn J, Charpentier C. Omicron SARS-CoV-2 variant: What we know and what we don’t. Anaesth Crit Care Pain Med. 2021;41(1):100998. doi:10.1016/j.accpm.2021.100998

